# Can Current Medical Examination Consumption Reduce Long-term Medical Expenses? - Analysis Based on CHARLS Data

**DOI:** 10.1101/2024.03.31.24305131

**Authors:** Juan Luo, Lulu San, Sunian Han, Liang Bi, Zhenpeng Ren

## Abstract

**Background:** With the implementation of the healthy China strategy, physical examination, a means of preventing diseases, has gradually been valued by health care department. Can health examination effectively alleviate the pressure on residents’ medical expenses?

**Method:** Based on the CHARLS2015 and CHARLS2018 databases, establishing an ordered multi-classification logistic regression model. To study the impact of residents’ health examination on the level of long-term medical expenditure.

**Results:** The results show that adhere to health examination, long-term physical exercise and quit smoking and drinking can reduce residents’ medical expenses. The physical examination behavior of different populations was further discussed based on gender. The study found that the more physical examinations of male residents, the less medical expenses, while the number of physical examinations of women is not significant for reduce medical expenses.

**Conclusion:** Therefore, China should vigorously improve the enthusiasm of different residents to participate in health check-ups, give full play to health check-ups in disease prevention and to reduce individual’s medical expenses.

## 1. Introduction

The report of the 20th National Congress of the Communist Party of China pointed out that China has built the world ‘s largest social security system. With the rapid growth of the economy, the health status of residents has been improved. However, there are also problems needed to resolve, such as increasing population aging, chronic diseases for young people, and excessive medical expenses. According to the ‘2021 National Economic and Social Development Statistical Bulletin ‘, the number of elderly people who aged 60 and over in China accounts for 18.7 % of the total population, of which the number of elderly people aged 65 and over accounts for 13.5% of the total population, far exceeding the UN ‘s 7% division standard. The degree of aging has been very serious in China. With the change of disease spectrum, chronic diseases have become the main cause of residents’ health problem. Chronic diseases in China show a trend of high incidence rate and younger age. ‘Health Management Blue Book: China Health Management and Health Industry Development Report (2018) ‘pointed out that the number of chronic diseases in China is about 300 million, of which 50% are under 65 years old. The proportion of death due to chronic diseases in urban and rural areas in China is as high as 85.3% and 79.5% respectively. Chronic disease treatment cycle is generally longer, residents need to pay higher medical costs.

In order to effectively reduce the high incidence rate and younger trend of chronic diseases in China and lowing the burden of residents ‘medical expenses, since 2016, the state has successively issued’ Healthy China 2030 ‘Planning Outline’ and ‘13th Five-Year’ Health and Health Plan’. The report of the 20th National Congress of the Communist Party of China further proposed to ‘put the protection of people’s health in a strategic position of priority development and improve people’s health promotion policies’. This fully reflects our party put people’s health and well-being in the first place and strive to create a high-quality national health care system. The World Health Organization has pointed out that through health check-ups, humans can effectively control and treat one-third of diseases, and studies have shown that for every additional yuan invested in health check-ups, medical expenses can be reduced by 4 to 7 yuan. Thus, encouraging residents to actively participate in health check-ups will undoubtedly become an effective means of preventing disease and alleviate the pressure on residents’ medical expenses.

## 2. Literature review

On the importance of health examination for early prevention of disease analysis, Wu Feiyan et al. (2014) through the analysis of the second people’s hospital of Yunnan province physical examination center from 2011 to 2013 data found that the positive rate of physical examination results as high as 75%, some of the top detection rate of disease, people in daily life generally will not have a strange feeling, but if there are complications, it will bring serious harm to life. Tang Wentao et al. (2019) discussed the relationship between disease delay and medical burden and concluded that the increase of delay time was significantly correlated with the increase of monthly medical expenses after diagnosis. Therefore, if the delayed population can be fully timely treatment, can effectively save the country’s medical costs. Wang (2017) analyzed the physical examination data of Chengdu Aerospace Hospital in 2015, and found that the detection rate of chronic diseases such as hyperlipidemia and hypertension was high, and further analyzed the main reasons for the high positive rate. At present, due to the influence of physiological, psychological factors and bad living habits, coupled with the increasingly serious environmental pollution, the threat to human health is increasing, and the risk of people suffering from various diseases is gradually increasing. Through regular physical examination, the risk of diseases in the body can be found as early as possible. For the diseases that have been suffered, it can also be found in time and effective treatment measures can be taken to minimize the disease. Shen Shuguang (2018) believed that medical insurance should also reflect the concept of prevention and health management, shift from focusing on disease treatment to focusing on disease management, increase the level of protection of preventive health care and ‘minor diseases’, and practice the basic concepts of ‘healthy China construction’ and ‘prevention first’. He Wen, Shen Shuguang (2018) The reasonable level of medical insurance outpatient security can play a certain role in practicing the concept of prevention, improving the level of health security, improving the efficiency of fund use, reducing moral hazard, and promoting hierarchical diagnosis and treatment. Yao Zelin (2016) left many patients with ‘minor diseases’ in the primary outpatient department, which can alleviate the pressure of large hospitals and promote the rational allocation of medical resources. At the same time, it can also reduce the pressure of patients’ difficult and expensive medical treatment and improve their health. However, health check-ups have long been neglected in some populations, Zhang et al. (2016) used data from the China Health Services Household Survey in Hunan Province to find that routine health checkups for hypertension and diabetes in young people aged 18-44 were underestimated.

On the study of the relationship between health examination and medical expenses, Zhang Aihua et al. (2014) conducted a comparative analysis of the medical expenses of medical examinees and non-medical examinees in the basic medical insurance for urban employees. It was found that the medical insurance for urban employees improved the utilization rate of physical examination and the overall health level. The hospitalization expenses of medical examinees were significantly lower than those of non-medical examinees. Health examination can alleviate the excessive growth of medical insurance expenses. In view of the deepening of China ‘s aging population and the reality of a substantial increase in medical expenses for the elderly, Hong Na (2012) dynamically simulated and calculated the medical expenses of the elderly population aged 60 and over who regularly participated in physical examination and less participated in physical examination. In the future, if the medical expenses after the popularization of physical examination in the whole society will be reduced by nearly 40% compared with the non-popular situation. Zhang et al. (2016), in a survey of the willingness of adult residents in Tianjin to participate in health examinations, found that when medical insurance did not reimburse the cost of physical examination, only less than 30% of residents were willing to participate; and nearly 70% of the residents in the medical insurance reimbursement part of the cost, willing to participate in health examination. Miao Yanqing et al. (2020) calculated the input and output of the major factors and their corresponding inspection items. The results showed that for every CNY 1 invested in the preventive health examination project of chronic diseases, it can save about 11.72-14.27 CNY direct medical expenses. On the contrary, some scholars have concluded that Cui Yujie et al. (2018) ‘s health examination will give residents health signals, which will show a significant increase in medical expenses and medical service utilization in a short period of time.

In summary, many scholars have put forward different views on whether physical examination can reduce medical expenses, but most of them focus on the perspective of early prevention of diseases and believe that early treatment and early screening of certain diseases can reduce the medical expenses of residents. This is fully in line with the development of China’s concept of big health, in the healthy development of China’s environment, the concept of ‘prevention first’ gradually developed, China’s medical insurance costs to reduce the payment of a new perspective: on the one hand that increase the ‘small disease’ treatment can effectively reduce the ‘big disease’ brought about by the high medical costs [1]; on the other hand, prevention and treatment of certain chronic diseases for residents, improve the health literacy of the whole people, and reduce medical expenses [2]. This led to a series of preventive medicine research. As an early means of prevention, physical examination can screen ‘minor diseases’ and reduce the medical expenses of residents [3]. Therefore, this study starts from the factors of physical examination to study the effect of physical examination on the payment of medical expenses.

## 3. Data sources and research methods

### 3.1 Data sources

The data studied in this paper are from the national baseline tracking survey conducted by the China Health and Retirement Longitudinal Study (CHARLS) in 2015 and 2018. CHARLS is a survey of Chinese middle-aged and elderly families and individuals. The respondents are randomly selected people aged 45 and above in the family. The CHARLS project began a national baseline survey in 2011 and tracked the samples every two years. The CHARLS data is currently updated to 2018. To ensure the representativeness of the sample, the CHARLS baseline survey covered 450 villages in 150 counties and districts across the country. By the time of the national follow-up in 2018, the sample had covered 23,000 respondents in a total of 12,400 households, representing the middle-aged and elderly population in China as a whole.

In this study, health examination was used as the core explanatory variable to screen whether the CHARLS questionnaire in 2015 and the CHARLS in 2018 participated in the physical examination. The influencing factors of medical expenses payment level were analyzed through the physical examination status of different groups in different periods. Through model comparison, the influence of current physical examination on the current medical expenses’ payment level and the influence of current physical examination on the long-term medical expenses’ payment level were analyzed. To explore the long-term cost control effect of health examination on medical insurance payment.

The paper takes whether to participate in health examination as the core explanatory variable. By matching, merging and screening the respondents who participated in health examination in 2015 and 2018 and those who did not participate in health examination, a total of 874 valid samples were screened. Among them, 374 people participated in health examination in both surveys and 500 people did not participate in health examination. The descriptive statistics of the samples are shown in table 1.

**Table 1.** Basic statistical information of samples.

| Feature | Classification | Frequency | Percentage |
| --- | --- | --- | --- |
| Gender | male | 783 | 89.6% |
|  | female | 91 | 10.4% |
| Age | $45 \leq X < 60$ years | 191 | 21.9% |
| | $X \geq 60$ years old | 683 | 78.1% |
| Marital status | Married | 748 | 85.6% |
|  | Unmarried / divorced / widowed | 126 | 14.4% |
| Health status | Good / Very Good / General Health | 629 | 72.0% |
|  | Not good / very bad | 245 | 28.0% |
| Whether suffering from chronic diseases | Yes | 737 | 84.3% |
|  | No | 137 | 15.7% |
| Whether disabled | Yes | 33 | 3.8% |
|  | No | 841 | 96.2% |
| Do you have cancer | Yes | 8 | 0.9% |
|  | No | 866 | 99.1% |
| Do you have cancer | Yes | 265 | 30.3% |
|  | No | 609 | 69.7% |
| Do you control the frequency of drinking | Yes | 406 | 46.5% |
|  | No | 468 | 53.5% |
| Whether physical exercise | Regular exercise | 711 | 81.4% |
|  | Little / never physical exercise | 163 | 18.6% |
| Whether physical examination | Two medical examinations | 374 | 42.8% |
|  | No medical examination twice | 500 | 57.2% |

### 3.2 Variable selection and explanation

#### 3.2.1 Explained variables

The research topic of this paper is whether the current physical examination can reduce the medical expenses, so the dependent variable of the study is the question of GE010-6 in the CHARLS database income, expenditure and asset questionnaire: ‘This questionnaire wants to understand the different types of consumption expenditures in your home, including direct medical expenses and indirect medical expenses’. Due to the left skewed distribution of the sample data, the virtual variable is used to calculate the discrimination of the test questions by using the principle of high and low grouping method in educational statistics. The medical expenses are sorted from low to high, and the medical expenses are calculated respectively. The critical values of 27% and 73% are divided into three levels: low, medium and high. Therefore, the dependent variable is an ordered multi-classification variable. Among them, the low-level Y=1, the medium level Y=2, and the high-level Y=3.After descriptive statistics on the level of medical expenses, it was found that the average level of medical expenses was 1.53, and the standard deviation was 0.78.

#### 3.2.2 Explanatory variables

The main explanatory variables of this paper are the preventive measures taken by residents. Specifically for the CHARLS database questionnaire whether physical exercise: ‘DA05 Do you usually have a continuous physical exercise activity for ten minutes per week?’, for categorical variables, regular exercise=1, little/never exercise = 2; did you quit smoking: ‘DA061 Are you smoking or quitting now?’, classified variable, smoking cessation = 1, smoking = 2; whether to control alcohol; ‘DA069 Are you reducing your drinking frequency for health now?’, is a categorical variable, minimize/control drinking frequency=1, do not control/often drink=2; whether health examination; ‘C001 When was your last physical examination?’ is a categorical variable, in the past two years to participate in physical examination=1, no physical examination= 2. These four independent variables are the main variables in this study, measuring the relationship between prevention and medical costs. There are also some factors that may affect residents’ medical insurance expenses, such as gender, age, marital status in the basic information questionnaire, self-health cognition in the health status and function questionnaire, whether suffering from chronic diseases, disability, cancer and other information are also included in the independent variables.

The specific variables and their assignments are shown in Table 2.

**Table 2.** Meaning and basic statistical characteristics of explanatory variables.

| Type | Explaining variables | Explain the meaning of variables | mean value | standard deviation |
| --- | --- | --- | --- | --- |
| Personal characteristics | Gender(X1) | The gender information of the respondents is represented as a categorical variable, where male=1, female=2 | 1.1 | 0.306 |
|  | Age(X2) | represents the age of the respondent, is a categorical variable, where 45 to 60 years old=1, over 60 years old=2 | 1.78 | 0.413 |
|  | Marital status(X3) | It indicates the marital status of the respondents, which is a categorical variable. Married living with spouse/married but not living with spouse for work and other reasons=1, divorced / widowed / never married=2 | 1.14 | 0.351 |
|  | Self-health cognition(X4) | Indicates self-rated health status of the respondent, is a categorical variable, good /good/fair=1, bad /very bad=2 | 1.28 | 0.449 |
| Health characteristics | Chronic disease(X5) | It represents the status of chronic diseases of the respondents, which is a categorical variable, yes=1, no=2 | 1.16 | 0.364 |
|  | Whether disabled(X6) | Represents the disability status of the respondents, is a categorical variable, yes=1, no=2 | 1.96 | 0.191 |
| Health<br>behavior | Do you have<br>cancer(X7) | represents the situation of the<br>respondents suffering from<br>cancer, is a categorical<br>variable, yes=1, no=2 | 1.99 | 0.095 |
|  | Whether to quit<br>smoking(X8) | Indicates whether the<br>respondent quit smoking for<br>health, is a categorical<br>variable, yes=1, no=2 | 1.3 | 0.46 |
|  | Do you control<br>the frequency<br>of drinking(X9) | Indicates whether the<br>respondent drinks less in<br>order to control the frequency<br>of drinking for health, is a<br>categorical variable, yes=1,<br>no=2 | 1.46 | 0.499 |
|  | Whether<br>physical<br>exercise(X10) | Indicates the participant's<br>daily exercise, classified<br>variable, regular exercise=1,<br>little/never exercise=2 | 1.19 | 0.39 |
|  | Whether to<br>participate in<br>physical<br>examination(X<br>11) | Indicates the physical<br>examination of the respondent<br>in two surveys, is a<br>categorical variable, physical<br>examination in both surveys=<br>1, no physical examination in<br>both surveys=2 | 1.57 | 0.495 |

#### 3.2.3 Research methods and models

According to the nature of the variable, the explanatory variable is a multi-category ordered variable, and the following ordered multi-category Logistic regression model is established:

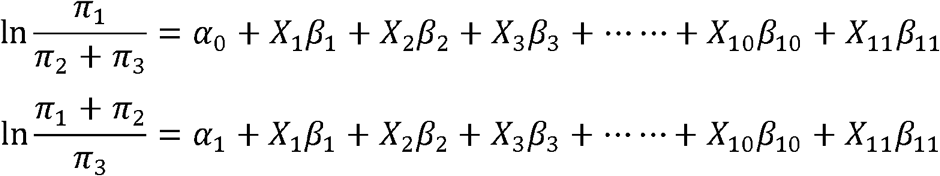

Among them, *α*_0_ and *α*_1_ are intercepts, respectively, *β*_*i*_ is the regression coefficient, write *π*_*i*_=P(Y=i) for medical expenses belong to different levels of probability, 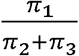 and 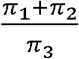 are the odds ratio (Odds Ratio, OR), take the natural logarithm.

## 4. Results analysis

### 4.1 Model fitting information test results

According to the above characteristics of variables, first of all, the model as a whole to fit information, parallel line hypothesis test. As shown in Table 3, the results show that the P value is less than 0.001, indicating that the model fitting results are better. In addition, as shown in Table 4, the parallel line hypothesis test result P is 0.178 greater than 0.05, and the original hypothesis is not rejected, indicating that the ordered multi-classification logistic regression model is suitable for analysis.

**Table 3.** Model fitting information.

| Model | -2 log likelihood | Chi-square | freedom | significance |
| --- | --- | --- | --- | --- |
| Intercept only | 721.506 |  |  |  |
| Finally | 546.86 | 174.646 | 11 | 0 |

**Table 4.** Parallel line test.

| Model | -2 log likelihood | Chi-square | freedom | significance |
| --- | --- | --- | --- | --- |
| Original hypothesis | 546.86 |  |  |  |
| Routine | 531.756 | 15.104 | 11 | 0.178 |

### 4.2 Analysis of Logistic model output results

By using spss23.0 software to analyze the model, the output results are shown in Table 5.

**Table 5.**
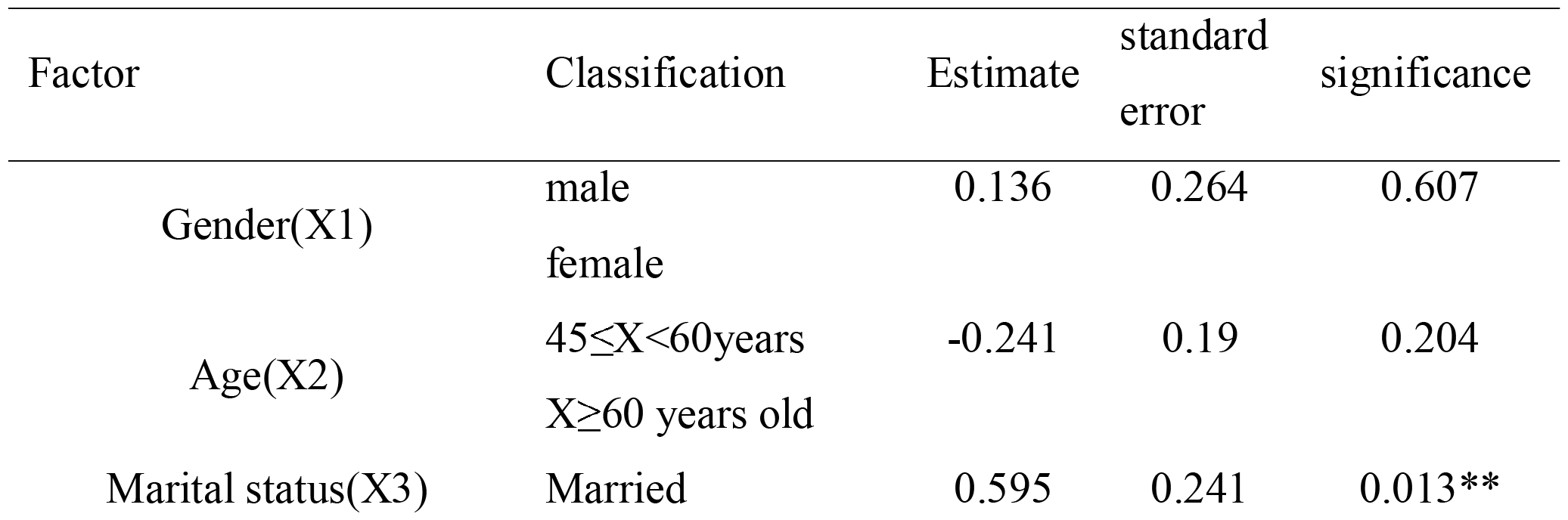
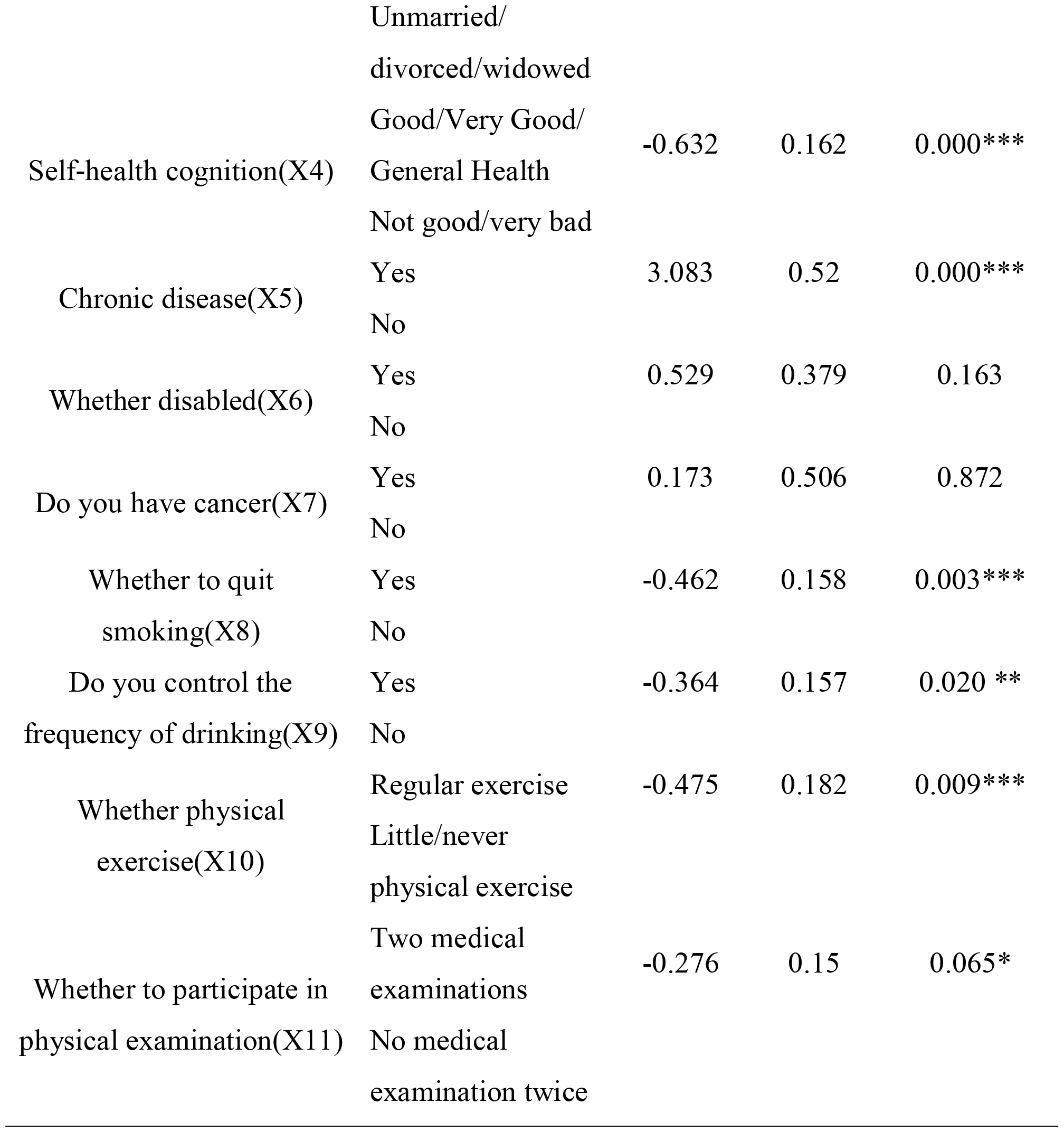
Logistics analysis of related variables.

The results show that marital status in personal characteristics information, health status in health characteristics information, whether suffering from chronic diseases, whether quitting smoking in health behavior, alcohol control, whether doing physical exercise in prevention information, whether participating in health check-ups have significant impact on the level of long-term medical expenditure, while gender, age, disability, whether suffering from cancer do not pass the significance test.

#### 4.2.1 Medical examination has a long-term control effect on the level of medical expenditure

The paper focuses on the relationship between preventive behavior and the level of medical expenses. Specifically, there is a significant relationship between the factors of continuous physical examination and the level of medical expenses (p< 0.1). The partial regression coefficient of physical examination in both surveys is-0.276, and the corresponding OR value is exp (-0.276) =0.76<1, indicating that the factors of physical examination are negatively correlated with the level of medical expenses. Compared with people who have not undergone physical examination, the possibility of a lower level of medical expenses for people undergoing physical examination is 0.76 times. That is to say, the level of medical expenses paid by the people who participated in the physical examination continuously in the two surveys is lower than that of the people who never participated in the physical examination. This is because the residents who participated in the physical examination continuously can achieve early detection and early treatment in the early stage of the disease, reducing the possibility of residents suffering from major diseases and thus reducing the medical expenses. Expenditure, and people who have never participated in physical examination, no investment in prevention, nor regular physical examination, their risk of major diseases is much higher than that of regular physical examination, so the level of medical expenses paid by people who have not undergone physical examination is higher than that of physical examination.

#### 4.2.2 Long-term physical exercise can reduce residents’ medical costs

Whether there is a significant relationship between physical exercise factors and the level of medical expenses payment p<0.001, the partial regression coefficient of people who exercise regularly is-0.475, and the corresponding OR value is exp (-0.475) =0.62<1, indicating that people who exercise regularly are negatively correlated with the level of medical expenses. Compared with people who do not exercise regularly, the possibility that the level of medical expenses of people who exercise regularly is one level lower is 0.62 times, indicating that the level of medical expenses of people who exercise regularly is lower than that of people who do not participate in physical exercise. This is because people who exercise regularly have enhanced physical fitness and improved immunity. Reduce the number of medical treatments, so its low level of medical expenses paid. That is, regular physical exercise and other physical activities can reduce medical expenses. In recent years, the state has issued many policies to support the development of the deep integration of sports and medical care, and actively implement non-medical health behavior intervention. Relevant policies have integrated physical exercise into disease and health management, and physical exercise has gradually integrated medical treatment, becoming the main low-cost preventive behavior to ensure the health of the whole people. In addition, the healthy China planning outline put forward the ‘combination of medicine and sports’, physical exercise is also a preventive behavior, preventive behavior affects the level of medical expenses.

#### 4.2.3 Whether to quit smoking and whether to control the frequency of drinking significantly affect the level of medical expenses

Among the health behavior variables, the p<0.001 of smoking cessation factor was significant at the 1 % level of medical expenses payment. The partial regression coefficient of smoking cessation factor was -0.462, and the corresponding OR value was exp (-0.462) =0.63<1, indicating that smoking cessation factor was negatively correlated with the level of medical expenses payment. Compared with smokers, smokers who quit smoking were 0.63 times more likely to pay a lower level of medical expenses. It shows that the level of medical expenses paid by people who quit smoking is lower than that of people who smoke. This is because people who quit smoking have a lower risk of certain diseases, such as lung airway disease and lung cancer, while people who smoke have an increased risk of these diseases. The factor of whether or not to control the frequency of drinking is significant at the level of 5 %, and its partial regression coefficient is -0.364, the corresponding OR value is exp (-0.364)=0.69<1, indicating that the factor of whether or not to control the frequency of drinking is negatively correlated with the level of medical expenses, and that people who control the frequency of drinking are 0.69 times more likely to pay a lower level of medical expenses than those who often drink. That is to say, the medical expenses of people who control drinking are lower than those of people who often drink, because people who control the frequency of drinking reduce the harm of alcohol to the body, while people who often drink increase the harm of alcohol to the body, so people who control the frequency of drinking have a low level of medical expenses, and people who often drink have a high level of medical expenses.

### 4.3 Gender-based heterogeneity analysis

To explore whether the impact of physical examination on the level of medical expenses payment in different populations is different, the paper analyzes the heterogeneity of gender factors in residents’ personal characteristics. The heterogeneity regression analysis model of gender characteristics is as follows. Model 1 and model 2 in the table, as shown in table 6.

**Table 6.** Regression results based on gender heterogeneity analysis.

| Variable | Model 1 | Model 2 |
| --- | --- | --- |

|  | male | female |
| --- | --- | --- |
| Gender | - | - |
|  | - | - |
| Marital status | -0.174**<br>(-2.54) | -0.146<br>(-0.89) |
| Health status | 0.268***<br>-4.66 | 0.158<br>-0.92 |
| Whether disabled | -0.203<br>(-1.49) | -<br>- |
| Whether physical exercise | -0.142***<br>(-2.70) | -0.003<br>(-0.02) |
| Whether to quit smoking | -0.138**<br>-2.52 | -0.046<br>-0.26 |
| Do you control the frequency of drinking | -0.094*<br>-1.88 | -0.099<br>-0.51 |
| Whether physical examination | -0.136***<br>(-2.76) | 0.108<br>-0.66 |
| Control variables | Yes | Yes |
| Sample size | 783 | 91 |
| Pseudo.R2 | 0.059 | 0.025 |

#### 4.3.1 Preventive behavior in the male sample has a significant effect on the level of medical expenses paid, while the female sample has no significant effect on the relationship

From the perspective of residents’ preventive behavior, the physical examination behavior of the male group is significant at the level of 1% of the medical expenses’ payment level, and its partial regression coefficient is -0.136, indicating that male residents invest in physical examination. The level of medical expenses payment will be reduced, while the female sample is not significant. From the analysis of the factors of physical exercise, the partial regression coefficient of the male group in the factors of physical exercise is negative, and the level of medical expenses payment is significant at the level of 1%. It shows that the people who often carry out physical exercise have a lower level of medical expenses payment. For the female group, whether to carry out daily exercise. Although its regression coefficient is negative, but not significant. It can be seen that the prevention behavior is significant in the male group and not significant in the female group. This may be due to the fact that the female group is more concerned about their health status in daily life than the male group, and the health behavior and health literacy of different gender groups vary due to different living habits[4].In terms of the relationship between gender and health literacy and medical expenses, Liang Keke and Song Zhijing (2021) studied the influencing factors of health knowledge and behavior literacy of residents in a district of Lanzhou City[5]. Through empirical data, it is concluded that women ‘s health behavior literacy level is higher than that of men. The reason is that men have a high rate of exposure to tobacco and alcohol, and it is not easy to adhere to healthy behavior, while women often take on the task of taking care of family life in the family, so they pay more attention to the concept of healthy life. Women will invest more health behaviors or preventive behaviors in their daily lives, resulting in that the physical examination behavior of female samples has little effect on the level of medical expenses payment, while men generally pay less attention to their own health status in daily life, or have poor health awareness. Smoking, drinking, staying up late and other behaviors will cause the occurrence of certain diseases, thus increasing the number of medical visits and medical expenses of men. When taking physical examination behavior, some potential diseases will be killed in the early stage of the disease, thus reducing the level of medical expenses payment. Ao Qin (2018) studied the influencing factors of health behavior literacy [6]. Health behavior literacy showed different distribution in gender, age and occupation, among which women ‘s health literacy was higher than that of men. Therefore, there are gender differences in preventive behavior. Women ‘s health behavior literacy is higher than that of men. Therefore, community publicity should be carried out in the context of national health to improve men ‘s health literacy and prevention awareness.

#### 4.3.2 Health behavior in the male sample significantly affected the level of medical expenses paid, while the female sample was not significant

Whether the residents quit smoking behavior is also different between the male sample and the female sample. Whether to quit smoking is significant at a 5 % significant level in the male sample, but not significant in the female sample. The partial regression coefficient is negative, indicating that male residents actively adopt smoking cessation behavior, the lower the level of medical expenses paid. The significance of whether residents control the frequency of drinking in male samples and female samples is also different. In the male sample, whether the frequency of drinking is controlled at a significant level of 10 % is significant, and the coefficient is negative, indicating that residents actively reduce drinking behavior. The lower the level of medical expenses paid, and the coefficient of the female sample is also negative, but not significant. The reason is also easy to understand, because most of the groups with smoking and drinking behavior are men. These bad living habits will lead to the occurrence of diseases. By investing in physical examination behavior, the potential risk factors of certain diseases are screened out in the early stage of disease evolution, which reduces the risk of some major diseases and also reduces the level of medical expenses paid by residents. Many scholars in China have similar conclusions. Some scholars have studied the relationship between smoking and drinking and the age of different genders [7-8], that is, the age of chronic diseases for men who had smoke and drink will be about 3 years earlier, while the factors of quitting smoking and reducing the frequency of drinking are not significant for women. The reason is that there are fewer people smoking and drinking in the female group. It can be seen that if men keep health behaviors can effectively reduce the level of medical expenses, which is the same as our research results. Therefore, whether to quit smoking or control the frequency of drinking in the male sample has a significant impact on the level of medical expenses payment, while the female sample is not significant. Some scholars believe that smoking and drinking have different effects on people of different ages, with less impact on young people and greater impact on the elderly. Long-term smoking and drinking can cause multiple organ diseases such as heart and lung in the elderly, which in turn induces various chronic diseases such as hypertension and diabetes. Advocating a good lifestyle can reduce the risk of illness and reduce the prevalence of chronic diseases in China [9]. In daily life, men with bad habits are higher than women. Therefore, it is further proved that men’s health behavior significantly affects the level of medical expenses, while women’s health behavior is not significant.

## 5. Recommendations

Through the above analysis, we can find that health examination can reduce the level of residents’ medical expenses in the long run, but only 43.4% of the population participated in health examination in both surveys, and the enthusiasm of residents to participate in health examination for a long time needs to be further improved. On the other hand, due to differences in subsidy standards, physical examination items, the level of medical institutions, resulting in uneven effect of health examination. The demand for national health examination in China is urgent, and there is a great potential for market development. However, if effective and standardized organization and implementation cannot be carried out, the effectiveness of health examination in preventing diseases and reducing medical expenses cannot be fully utilized.

### 5.1 Strengthening publicity and education to improve residents’ enthusiasm for physical examination

The idea of ‘preventive treatment of disease’ in traditional Chinese medicine was first produced in the Spring and Autumn Period and the Warring States Period. In the Tang Dynasty, Sun Simiao had the warning of ‘preventive treatment of disease by doctors. It is necessary to vigorously promote the concept of preventive treatment of disease in the whole society. When there is no disease, it is necessary to do a good job in the prevention of disease. Grassroots communities and medical service institutions can organize health lectures, issue leaflets and other ways to carry out the publicity and education of health care knowledge of traditional Chinese medicine, as an effective supplement to the health examination mode of western medicine, to help the residents understand the risk factors affecting health. Develop good living habits while actively participating in health examinations.

### 5.2 Standardize inspection projects to ensure the effectiveness of work

May 10, 2014 ‘health examination basic project expert consensuses officially released. The basic item list of health examination is clear, covering physical examination, general examination and auxiliary examination. At present, there are three main modes of health examination institutions on the market. Professional physical examination institutions, physical examination centers affiliated to general hospitals and physical examination institutions attached to other industries should further standardize the examination items of different types of physical examination institutions. While giving full play to the professional and standardized characteristics of physical examination centers in public hospitals, we should encourage and guide the development of other types of physical examination institutions and promote effective benign competition. Physical examination organizations should do a good job in the relevant notification before the physical examination, to further improve the technical level, combined with the physical examination of their own actual situation, according to the results of physical examination to give effective assessment, to develop targeted health management recommendations and programs.

### 5.3 System integration, physical examination into the medical insurance system

In recent years, the voice of incorporating physical examination into medical insurance has been increasing. At the national and local meetings, members had also put forward relevant bills, and suggested that physical examination should be included in the scope of medical insurance reimbursement. The current medical insurance system in China does not include all kinds of medical expenses in the basic medical insurance diagnosis and treatment project. This is a continuation of the past public expense and labor insurance medical policy. It only divides the health examination into the general physical examination organized by the employers and the health examination at the expense of the individual for special purposes. In fact, localities managers have successively carried out health check-ups for urban residents’ medical insurance and new rural cooperative medical insurance contributors, and have also introduced corresponding policy measures, which have improved the participation in health check-ups in China to a certain extent and enhanced the health care awareness of urban and rural residents.

## Data availability statement

The data in this study can be found in the official website of China Health and Longitudinal Study, CHARLS:http://charls.pku.edu.cn/

## Author contributions

Juan Luo put forward the research ideas and framework of this paper.Juan Luo wrote the full text.Lulu Shan and Zhenpeng Ren data processing and analysis. Sunian Han and Liang Bi combed the literature and checked and revised the full text.All authors have read and agreed to the published version of the manuscript.

## Acknowledgement

[Fund Project] National Social Science Fund Key Project: Research on the Mechanism, Path and Countermeasures of Breaking the ‘Digital Divide’ of the Elderly (No.21AGL024)

## Ethics statement

This study does not involve other people’s privacy and does not violate ethics.

## References

[1] Wu Jing, Li Li, Li Yinghua. The level of chronic disease prevention literacy and its influencing factors among Chinese residents in 2016 [J]. China Health Education, 2018,34 (05): 22–26.

[2] Hakro, Saifullah; Jinshan, Li. Workplace Employees’ Annual Physical Checkup and During Hire on the Job to Increase Health-care Awareness Perception to Prevent Dise ase Risk: A Work for Policy-Implementable Option Globally[J]. Safety and Health at Work,2019(08):132–140

[3] Tang Wentao, Iwasaki Hongjie. Economic evaluation of the burden of delayed diagnosis and treatment of diabetes based on Japanese medical big data [J]. Chinese Journal of Evidence-based Medicine, 2019, 19 (04): 392–397.

[4] Cho, MK (Cho, Mi-Kyoung); Cho, YH (Cho, Yoon-Hee). Role of Perception, Health Beliefs, and Health Knowledge in Intentions to Receive Health Checkups among Young Adults in Korea[J]. International Journal of Environmental Research and Public Health, 2022(11):21.

[5] He Wen, Shen Shuguang. Can medical insurance “protect minor illness” shoulder health care and cost control [J]. Insurance Research, 2018 (11): 93–106.

[6] Sofue, T (Sofue, Tadashi); Hara, T (Hara, Taiga) et al. Changes in Prevalence and Health Checkup Coverage Rate of Chronic Kidney Disease (CKD) after Introduction of Prefecture-Wide CKD Initiative: Results of the Kagawa Association of CKD Initiatives[J]. Journal of Personalized Medicine, 2021(12):11

[7] Zhang L,Zhang W,Zhang L,et al. Associations of Undergoing a Routine Medical Examination or Not with Prevalence Rates of Hypertension and Diabetes Mellitus: A Cross-Sectional Study[J]. International Journal of Environmental Research & Public Health, 2016,13(7):628.

[8] Yu, WY (Yu, Wenya); Liu, X (Liu, Xiang)et al. Control of unreasonable growth of medical expenses in public hospitals in Shanghai, China: a multi-agent system model[J]. BMC Health Services Research, 2020(06):29

[9] Sase, Y (Sase, Yuji); Kumagai, D (Kumagai, Daiki)et al. Characteristics of Type-2 Diabetics Who are Prone to High-Cost Medical Care Expenses by Bayesian Network[J]. International Journal of Environmental Research and Public Health, 2020(09):23.

[10] Miao Yanqing, Zhang Bingli. Input-output analysis of preventive health examination for chronic diseases [J]. China Health Policy Research, 2020,13 (05): 19–25.

[11] Cui Yujie, Yao Yao, Liu Guoen, Yang Maorui. Has physical examination changed people ‘s medical behavior? An Analysis Based on NCMS Health Examination Data [J]. Insurance Research, 2018 (02): 53–64.

